# Age-related disparity of survival outcomes and treatment-related adverse events in patients with metastatic colorectal cancer

**DOI:** 10.1101/2022.10.08.22280865

**Authors:** Lingbin Meng, Ram Thapa, Maria G. Delgado, Maria F. Gomez, Rui Ji, Todd C. Knepper, Joleen M. Hubbard, Xuefeng Wang, Jennifer B. Permuth, Richard D. Kim, Damian A. Laber, Hao Xie

## Abstract

**Background:** While the incidence of newly diagnosed early-onset colorectal cancer has been increasing, age-related disparity of survival outcome and treatment-related adverse events in patients with metastatic CRC (mCRC) has been inadequately studied with inconclusive findings. In this study, we aimed to evaluate such age-related disparity in this patient population.

**Methods:** We used individual patient data from three clinical trials (Study 1: NCT00272051, NCT 00305188 and Study 2: NCT00364013) in Project Data Sphere. All patients were diagnosed with mCRC and received first-line 5-fluorouracil and oxaliplatin. Clinical and genomic data of 763 patients with mCRC from Moffitt Cancer Center were used to assess genomic alterations and serve as an external and real-world validation cohort to evaluate overall survival (OS) disparity. Survival outcomes and treatment-related adverse events were estimated and compared in patients among three age groups: <50, 50-65, and >65 years.

**Results:** Among 1223 patients from previous clinical trials, 179 (14.6%) were younger than 50 years. These patients had significantly shorter progression-free survival (PFS) (HR=1.46; 95%CI=1.22–1.76; *p*<0.001) and OS (HR=1.48; 95%CI=1.19–1.84; *p*<0.001) compared to patients in the 50-65 group of both Study 1 and Study 2 after adjustment for gender, race, and performance status. Significantly shorter OS was also observed in patients from the <50 group in the Moffitt cohort. When compared to other age groups, the <50 group had significantly higher incidence of nausea/vomiting (69.3% vs 57.6% vs 60.4%, *p*=0.019), severe abdominal pain (8.4% vs 3.4% vs 3.5%, *p*=0.018), severe anemia (6.1% vs 1.0% vs 1.5%, *p*<0.001), and severe rash (2.8% vs 1.2% vs 0.4%, *p*=0.047), but significantly lower incidence of fatigue, severe diarrhea, severe fatigue, and severe neutropenia. The <50 group had earlier onset of nausea/vomiting (1.0 vs 2.1 vs 2.6 weeks, *p*=0.012), mucositis (3.6 vs 5.1 vs 5.7 weeks, *p*=0.051), and neutropenia (8.0 vs 9.4 vs 8.4 weeks, *p*=0.043), and shorter duration of mucositis (0.6 vs 0.9 vs 1.0 weeks, *p*=0.006). In the <50 group, severe abdominal pain and severe liver toxicity were associated with both shorter OS and PFS. In contrast, moderate peripheral neuropathy was associated with longer PFS. Our genomic data showed that the <50 group had higher prevalence of *CTNNB1* mutation (6.6% vs 3.1% vs 2.3%, *p*=0.047), *ERBB2* amplification (5.1% vs 0.6% vs 2.3%, *p*=0.005), and *CREBBP* mutation (3.1% vs 0.9% vs 0.5%, *p*=0.050), but lower prevalence of *BRAF* mutation (7.7% vs 8.5% vs 16.7%, *p*=0.002).

**Conclusions:** Patients with early-onset mCRC had worse survival outcome and unique adverse-event patterns, which could be partially attributed to distinct genomic profiles. Our findings might improve an individualized approach to chemotherapy, counseling, and management of treatment-related adverse events in this patient population.

## Introduction

Colorectal cancer (CRC) is the third most common malignancy and the second leading cause of cancer-related death worldwide [1]. Despite the continuous decline of overall incidence and mortality of CRC in the United States as a result of nationwide-adopted screening program [2-4], the incidence of the early-onset CRC (EO-CRC), patients diagnosed with CRC before 50 years of age has been increasing by approximately 2% annually since the 1990s [5, 6]. It is projected that 10.9% of all colon cancers and 22.9% of all rectal cancers will be diagnosed as EO-CRC by 2030 [7, 8].

Neither the etiology of increased incidence nor the distinct biology of EO-CRC compared to its older counterparts are clearly revealed in the literature. For example, inherited cancer syndromes such as Lynch syndrome may be more prevalent in EO-CRC patients than in all patients with CRC. However, only 5-7% of the patients with EO-CRC carry those deleterious germline mutations of mismatch repair genes. Therefore, most EO-CRCs are sporadic in nature [9, 10]. Previous studies examining genomic profile of EO-CRC showed few molecular differences between EO-CRC and its older counterparts, except for an enrichment of microsatellite instability as expected from higher prevalence of Lynch syndrome in patients with EO-CRC [11]. Even the few molecular differences reported were often not confirmed in other studies likely due to selection bias in different patient cohorts [8, 12, 13]. Some studies investigated nongenetic risk factors for CRC, such as low physical activity, excess alcohol consumption, smoking, and obesity, but the results were neither conclusive nor adequately explaining the increased incidence of EO-CRC [14-16].

The increased incidence of EO-CRC poses unique challenges to the management of young patients with all stages of CRC, especially with regard to the prognosis and treatment-related adverse events [17]. In current clinical practice, patients with EO-CRC receive similar if not more aggressive therapies in the advanced disease setting compared to their older counterparts. Fluoropyrimidine-based combination chemotherapy plus biologics remains the most common first-line therapy for patients with metastatic CRC (mCRC) [18]. Triplet chemotherapy combination of fluoropyrimidine, oxaliplatin, and irinotecan (FOLFOXIRI) is an alternative for patients who are younger, have more aggressive disease and good performance status, offering a better treatment outcome at the expense of worse toxicities [19]. Although both anti-vascular endothelial growth factor (VEGF) antibody [20, 21] and anti-epidermal growth factor receptor (EGFR) antibodies in combination with chemotherapy showed comparable benefit in patients with advanced CRC of all ages [22, 23], it is unclear whether there is disparity in treatment-related adverse events and outcome from advanced CRC between early-onset and its older counterparts. Previous studies suggested an increased incidence of some specific toxicities such as nausea/vomiting in early-onset mCRC (EO-mCRC), but the data was largely limited in the adjuvant setting and findings across studies were not always in agreement [24-28]. Similarly, some studies reported a poorer survival in patients with advanced EO-CRC [24, 25, 29], whereas others observed a similar survival outcome between early-onset and average-onset patients with mCRC [26-28, 30, 31]. These discrepancies might be due to selection bias and/or the inclusion of older patients (age>65) in the average-onset group for comparison.

To address this literature gap, we used individual patient data from three clinical trials where patients with mCRC received first-line 5-fluorouracil and oxaliplatin (FOLFOX), and a real-world patient cohort to compare the differences of treatment outcome and adverse events among three different age groups of patients with mCRC.

## Patients and Methods

We used individual patient data from three multi-center, randomized phase III clinical trials in the Project Data Sphere and grouped them into Study 1 (NCT00272051, NCT00305188) and Study 2 (NCT00364013). Clinical trials in Study 1 evaluated the efficacy of Xaliproden in reducing (NCT00272051) or preventing (NCT 00305188) the neurotoxicity of FOLFOX as first-line treatment for patients with mCRC. The clinical trial in Study 2 evaluated the efficacy of panitumumab in combination with FOLFOX as first-line therapy for patients with mCRC. Only individual patient data from the control (FOLFOX only) arms of the three clinical trials were combined and used in our study. Study design, inclusion and exclusion criteria, interventions, endpoints, adverse events, and outcome of the three clinical trials have been previously reported [23, 32-35]. Given these patients were treated in a clinical trial setting prior to 2010, we included a real-word patient population as an external validation cohort for overall survival (OS). Patients of this cohort were identified from prospectively maintained Moffitt Clinical Genomic Action Committee Database and CARIS clinical database. These include patients diagnosed with mCRC and treated at Moffitt Cancer Center (MCC) from 2006 to 2022 and have available clinical and next-generation sequencing (NGS) data. Clinical NGS data of these patients were provided by common commercially available platforms including FoundationOne, FoundationOne ACT, FoundationOne CDx, CARIS, and Guardant360 along with an in-house NGS assay referred to as Moffitt STAR. These platforms have been described in depth elsewhere previously [36-38]. This study was approved by the Institutional Review Board by MCC through Advarra.

OS was defined as the time from randomization to death. Participants who were alive at the analysis data cutoff were censored at their last contact date. Progression-free survival (PFS) was defined as the time from randomization to date of disease progression assessed radiographically per the Response Evaluation Criteria in Solid Tumors (RECIST) 1.0. Safety endpoints included incidence of any grade and grade 3-5 (severe) treatment-related adverse events according to the NCI-CTCAE version 3.0, the time of their onset, and their duration. Analyses of the differences of genetic alterations among three age groups were exploratory in nature.

All patients were divided into 3 age groups: <50, 50-65, and >65 years. The χ^2^ test and Fisher’s exact test were used to test the distribution of baseline characteristics, adverse events, and NGS genomic features in the 3 groups. Continuous variables were compared with Kruskall-Wallis test and chi-squared or exact Fisher test were performed for categorical variables. OS and PFS were evaluated according to the Kaplan-Meier method and compared using the log-rank test. Cox proportional hazards model was used to determine the association of adverse events with OS and PFS. P values in this study were 2-sided, and a *P* value less than 0.05 was considered statistically significant. Pairwise comparisons adjusting for multiple testing were performed using the Benjamini&Hochberg method.

## Results

### Demographics and clinical characteristics of patients with mCRC

Among all 1959 patients included, 1223 were from Study 1 and Study 2 for analyses of survival outcomes and treatment-related adverse events. 736 patients were from the Moffitt cohort for assessment of OS and tumor genetic alterations. As shown in **Supplemental table 1**, the Moffitt cohort included a significantly higher rate of EO-mCRC patients (*p*<0.001), but lower prevalence of white patients (*p*<0.001). Of the patients from Study 1 and Study 2, 179 (14.6%), 582 (47.6%), and 462 (37.8%) patients were in age<50, 50-65, and >65 groups at the time of stage IV CRC diagnosis, respectively (**Table 1**). There were significantly more women in age<50 group in contrast to more men in age>65 group (*p*<0.001). Fewer white and more non-white individuals (*p*=0.012) were represented in age<50 group. No significant difference of Eastern Cooperative Oncology Group (ECOG) performance status was observed among the three age groups (*p*=0.32).

**Table 1.** Demographic and clinical characteristics of patients with metastatic colorectal cancer from Study 1 and Study 2 stratified by age groups.

|  | < 50<br>(n=179) | 50 - 65<br>(n=582) | > 65<br>(n=462) | P value | Overall<br>(n=1223) |
| --- | --- | --- | --- | --- | --- |
| Gender, n (%) |  |  |  | <0.001 |  |
| Female | 98 (54.7) | 226 (38.8) | 162 (35.1) |  | 486 (39.7) |
| Male | 81 (45.3) | 356 (61.2) | 300 (64.9) |  | 737 (60.3) |
| Race, n (%) |  |  |  | 0.012 |  |
| White | 158 (88.3) | 551 (94.7) | 431 (93.3) |  | 1140 (93.2) |
| Other | 21 (11.7) | 31 (5.3) | 31 (6.7) |  | 83 (6.8) |
| ECOG, n (%) |  |  |  | 0.32 |  |
| 0/1 | 175 (98.3) | 565 (97.1) | 442 (96.1) |  | 1182 (96.9) |
| 2 | 3 (1.7) | 17 (2.9) | 18 (3.9) |  | 38 (3.1) |

### Age-related disparity of survival outcome in patients with mCRC

In Study 1, patients in the age<50 group had shorter median OS compared to the age 50-65 and age>65 groups (15.5 vs 20.5 vs 20.8 months, *p*=0.0026, **Figure 1A**). Patients in the age<50 group also had shorter median PFS than that of the other two age groups (8.1 vs 9.4 vs 8.6 months, *p*=0.0044, **Figure 1B**). Similar findings were observed in Study 2 as shown in **Figure 1C** (median OS: 18.4 vs 22.7 vs 18.0 months, *p*=0.0078) and in **Figure 1D** (median PFS: 7.3 vs 9.3 vs 8.7 months, *p*=0.0048). In univariate survival analysis, age<50 years (age<50 vs age 50-65: HR=1.50; 95%CI=1.21–1.85; *p*<0.001) and poor ECOG performance status (2 vs 0/1: HR=2.49; 95%CI=1.75–3.54; *p*<0.001) were identified as poor prognostic factors for OS in both Study 1 and Study 2 combined. Age<50 years was confirmed in multivariable analysis as an independent poor prognostic factor for OS (age<50 vs age 50-65: HR=1.48; 95%CI=1.19–1.84; *p*<0.001) after adjustment for gender, race, and ECOG performance status. Age<50 years was similarly found to be an independent poor prognostic factor for PFS (age<50 vs age 50-65: HR=1.46; 95%CI=1.22–1.76; *p*<0.001) as shown in **Table 2**.

**Figure 1.**
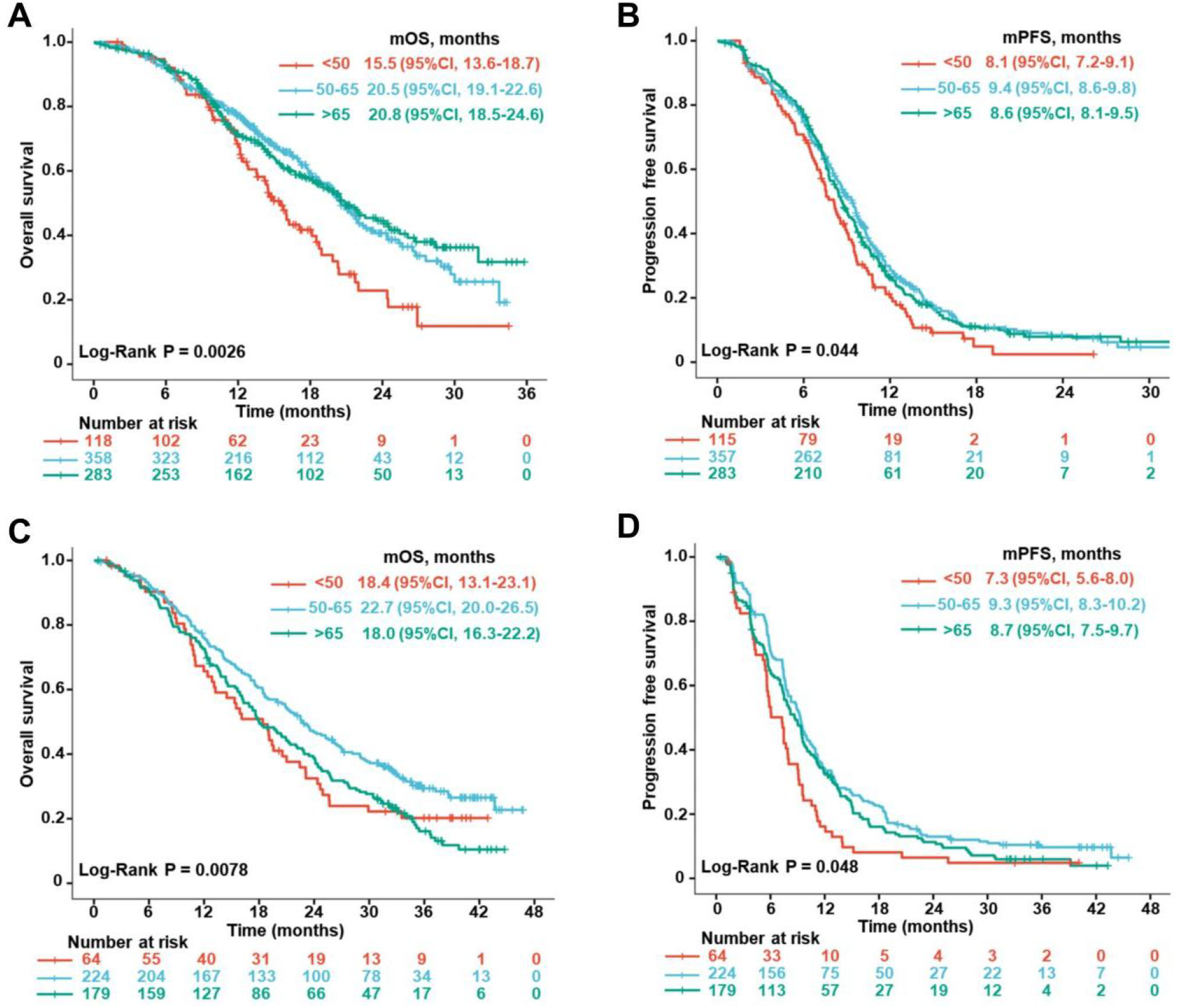
Survival outcome of patients with metastatic colorectal cancer stratified by age groups. (A) Overall survival and (B) Progression-free survival of patients with metastatic colorectal cancer from Study 1; (C) Overall survival and (D) Progression-free survival of patients with metastatic colorectal cancer from Study 2.

**Table 2.** The prognostic values of age groups and other common demographic and clinical factors in patients with metastatic colorectal cancer from Study 1 and Study 2.

| Overall survival |  |  |  |  |  |
| --- | --- | --- | --- | --- | --- |
| Variable |  | Univariate |  | Multivariable |  |
|  |  | HR (95%CI) | P value | HR (95%CI) | P value |
| Age | <50 vs 50-65 | 1.50 (1.21 - 1.85) | <0.001 | 1.48 (1.19 - 1.84) | <0.001 |
|  | >65 vs 50-65 | 1.17 (1.00 - 1.38) | 0.053 | 1.15 (0.97 - 1.35) | 0.10 |
| Gender | Male vs Female | 0.98 (0.84 - 1.14) | 0.82 | 1.00 (0.86 - 1.17) | 0.99 |
| Race | Other vs White | 1.19 (0.87 - 1.62) | 0.27 | 1.09 (0.80 - 1.49) | 0.57 |
| ECOG | 2 vs 0/1 | 2.49 (1.75, 3.54) | <0.001 | 2.41 (1.69 - 3.44) | <0.001 |
| Progression-free survival |  |  |  |  |  |
| Variable |  | Univariate |  | Multivariable |  |
|  |  | HR (95%CI) | P value | HR (95%CI) | P value |
| Age | <50 vs 50-65 | 1.46 (1.22 - 1.75) | <0.001 | 1.46 (1.22 - 1.76) | <0.001 |
|  | >65 vs 50-65 | 1.10 (0.97 - 1.26) | 0.15 | 1.10 (0.96 - 1.26) | 0.17 |
| Gender | Male vs Female | 0.98 (0.87 - 1.11) | 0.78 | 1.01 (0.89 - 1.14) | 0.92 |
| Race | Other vs White | 0.95 (0.74 - 1.22) | 0.69 | 0.89 (0.69 - 1.14) | 0.37 |
| ECOG | 2 vs 0/1 | 1.54 (1.10 - 2.15) | 0.011 | 1.55 (1.11 - 2.17) | 0.010 |

### Age-related disparity of treatment-related adverse-event pattern in patients with mCRC

Examination of treatment-related adverse events of all patients from Study 1 and Study 2 combined revealed a unique pattern for patients in the age<50 group (Figure 2). As shown in **Figure 2A, 2B**, and **Supplemental table 2**, compared to the other two age groups, the age<50 group had higher incidence of nausea/vomiting (69.3% vs 57.6% vs 60.4%, *p*=0.019), severe abdominal pain (8.4% vs 3.4% vs 3.5%, *p*=0.018), severe anemia (6.1% vs 1.0% vs 1.5%, *p*<0.001), severe rash (2.8% vs 1.2% vs 0.4%, *p*=0.047), but lower incidence of fatigue (44.1% vs 46.9% vs 55.6%, *p*=0.0052), neutropenia (38.5% vs 39.7% vs 49.8%, *p*=0.0018), severe diarrhea (6.1% vs 9.1% vs 13.0%, *p*=0.02), severe fatigue (4.5% vs 5.5% vs 9.5%, *p*=0.019), and severe neutropenia (25.7% vs 26.5% vs 38.1%, *p*<0.001). The age<50 group had earlier onset of nausea/vomiting (1.0 vs 2.1 vs 2.6 weeks, *p*=0.012), mucositis (3.6 vs 5.1 vs 5.7 weeks, *p*=0.051), and neutropenia (8.0 vs 9.4 vs 8.4 weeks, *p*= 0.043) as shown in Figure 2C and Supplemental table 3. Finally, the age<50 group had shorter duration of mucositis (0.6 vs 0.9 vs 1.0 weeks, *p*=0.006) as shown in **Figure 2D** and **Supplemental table 4**.

**Figure 2.**
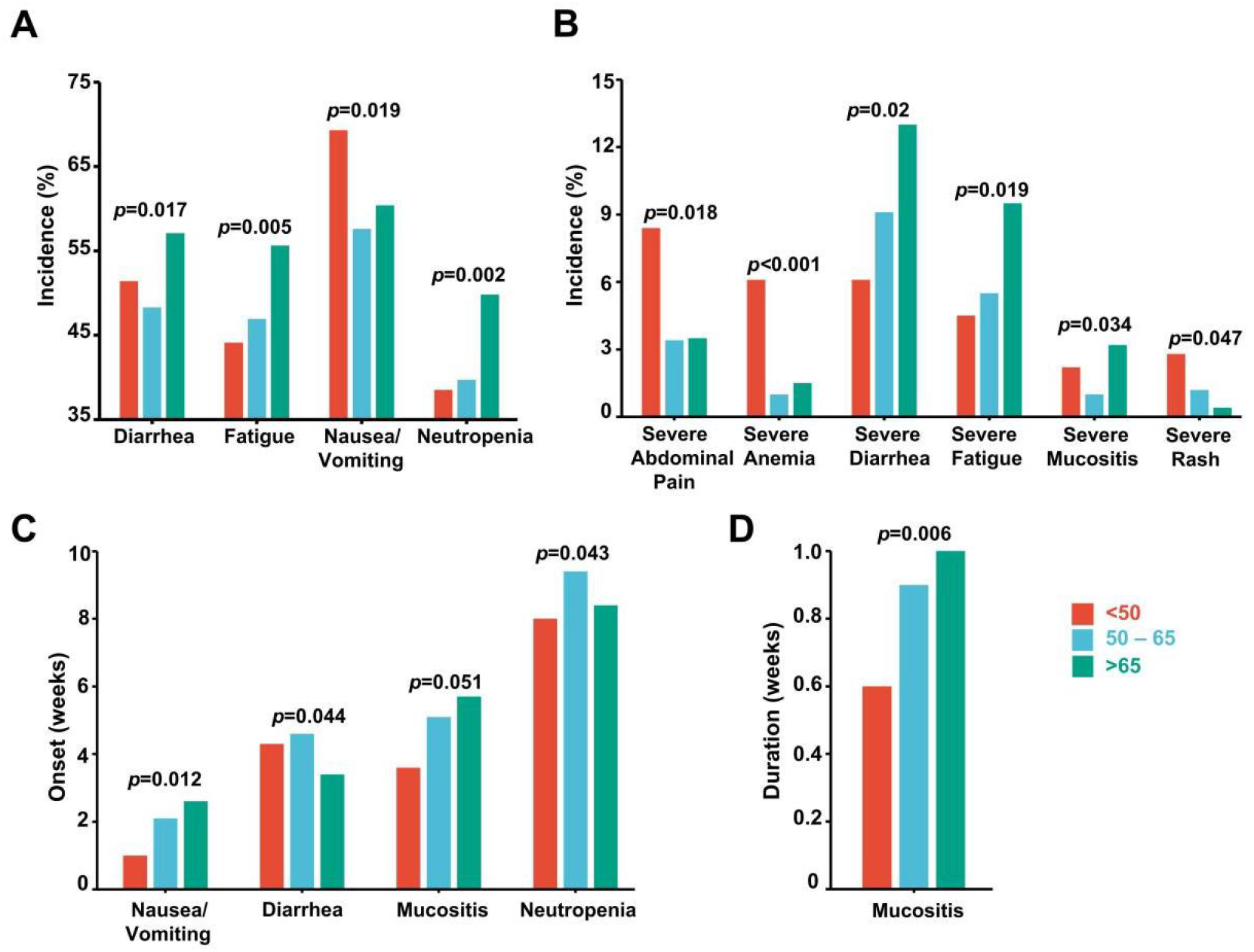
Adverse event pattern of patients with early-onset metastatic colorectal cancer in Study 1 and Study 2. (A) Incidence of adverse events; (B) Incidence of severe (grade 3-5) adverse events; (C) Median time to onset of adverse events; (D) Median time to resolution of adverse events for each age group with statistically significant differences.

### The prognostic value of treatment-related adverse events in patients with EO-mCRC

As summarized in **Figure 3A** and **Supplemental table 5**, severe abdominal pain (HR=2.24; 95%CI=1.23–4.09; *p*=0.008), severe live toxicity (HR=3.99; 95%CI=0.95–16.76; *p*=0.059) were poor prognostic factors for OS. In contrast, moderate (grade 1/2) fatigue (HR=0.66; 95%CI=0.44–0.97; *p*=0.008), moderate mucositis (HR=0.64; 95%CI=0.43–0.97; *p*=0.035), and moderate rash (HR=0.63; 95%CI=0.40–1.00; *p*=0.049) were prognostic factors for better OS. Similarly, severe abdominal pain (HR=2.51; 95%CI=1.43–4.41; *p*=0.001), severe live toxicity (HR=2.82; 95%CI=0.89–8.97; *p*=0.079) were poor prognostic factors for PFS. In contrast, moderate peripheral neuropathy (HR=0.43; 95%CI=0.26–0.71; *p*=0.001) was prognostic for better PFS (**Figure 3A** and **Supplemental table 5**).

**Figure 3.**
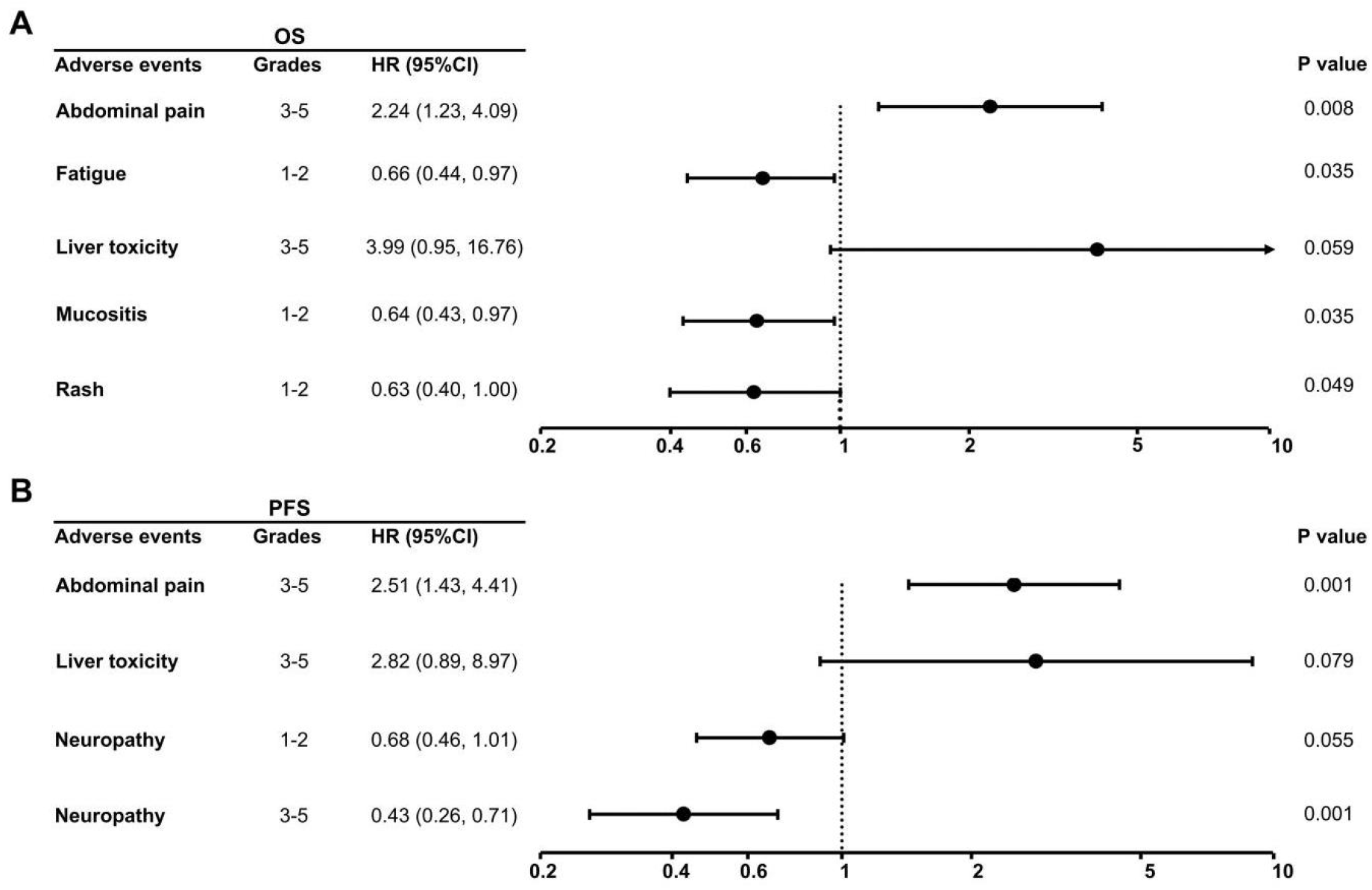
The prognostic value of adverse events in patients with metastatic colorectal cancer from Study 1 and Study 2. Hazard ratio of adverse events for (A) overall survival and (B) progression-free survival with statistical significance or a trend toward significance.

### Survival outcome and genetic alterations in patients with mCRC from the MCC cohort

As shown in Supplemental table 6, of the 736 patients in the MCC cohort, 196 (26.6%), 319 (43.3%), and 221 (30.1%) patients were in the age<50, age 50-65, and age>65 groups, respectively. The age<50 group were more frequent to receive triplet therapy (*p*<0.001) and carry left-sided tumor (*p*<0.001), while the age>65 group had a higher rate of white patients (*p*=0.044). Despite the difference of baseline characteristics between the MCC cohort and Study 1 and 2 (**Supplemental table 1**), patients in the age<50 group consistently showed worse median OS compared to the age 50-65 group and comparable median OS to the age>65 group (39.2 vs 51.3 vs 38.0 months, *p* = 0.018, **Figure 4A**). Further examination of genetic alterations was performed to explore the potential underlying causes of this age-related disparity of survival outcome in patients with mCRC. As listed in **Figure 4B** and **Supplemental table 7**, we found that all three age groups had very similar genetic alterations except that compared to the other two age groups, tumor of patients in the age<50 group had higher prevalence of *CTNNB1* mutation (6.6% vs 3.1% vs 2.3%, *p*=0.047), *ERBB2* amplification (5.1% vs 0.6% vs 2.3%, *p*=0.005), and *CREBBP* mutation (3.1% vs 0.9% vs 0.5%, *p*=0.050), but lower incidence of *BRAF* mutation (7.7% vs 8.5% vs 16.7%, *p*=0.002) as shown in **Figure 4B** and **Supplemental table 7**.

**Figure 4.**
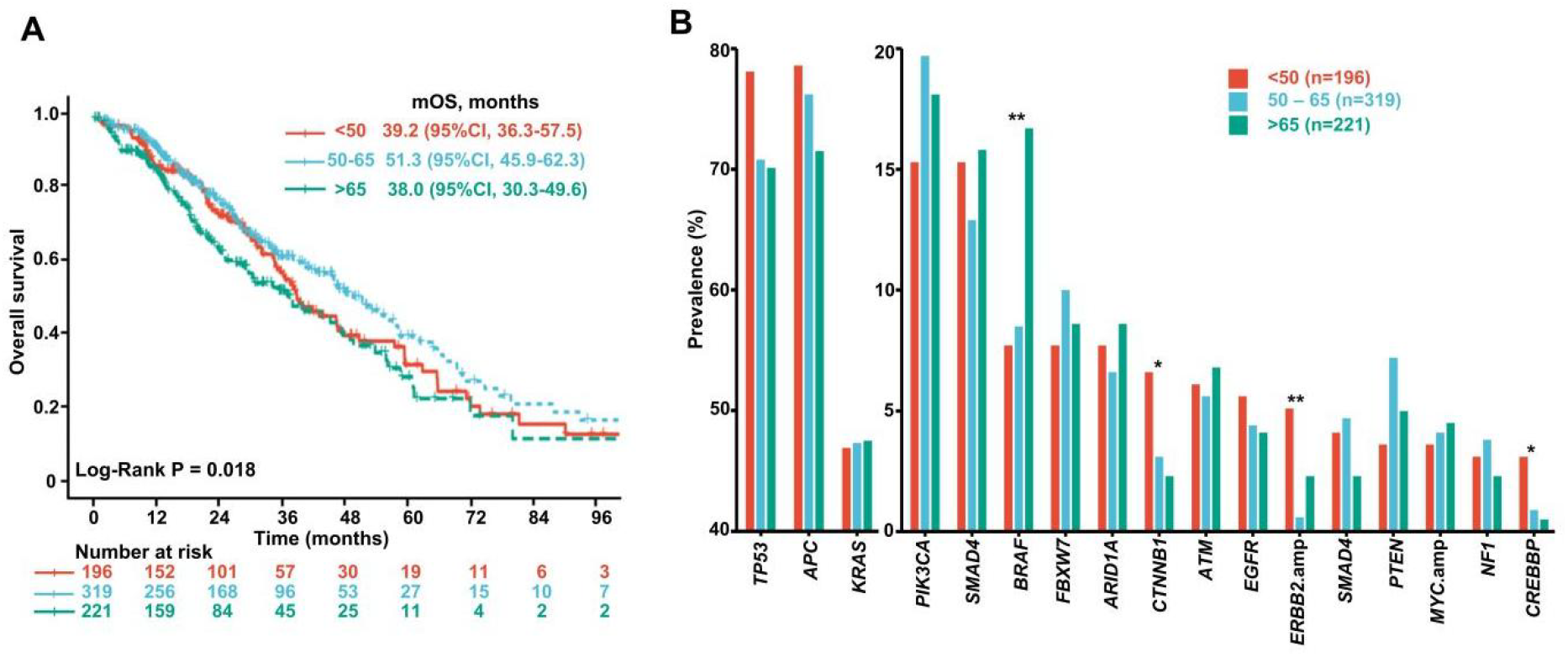
(A) Overall survival and (B) Prevalence of common gene mutations in patients with metastatic colorectal cancer from Moffitt Cancer Center cohort stratified by age groups. * *p* < 0.05; ** *p* < 0.01.

## Discussion

Hypotheses for increased incidence of EO-CRC despite a decline in overall incidence [5, 6] was that EO-CRC may be a different disease compared with its older counterparts as reflected by distinct clinicopathological and molecular features [5, 39]. Previous studies that attempted to investigate such age-related disparity of survival outcomes had discordant findings. Several studies found an association between younger age and worse survival [25, 29, 40, 41], whereas more studies revealed opposite findings [24, 26, 27, 30, 42, 43]. Lieu and colleagues observed younger and older ages were associated with worse survival in patients with mCRC [29]. However, all recent studies showed at least comparable [26-28, 30, 31] if not better prognosis [44] in patients with EO-mCRC. To the best of our knowledge, the present study is the first analysis of individual patients data from clinical trials and a more contemporary real-world cohort that demonstrated worse survival outcomes in patients with EO-mCRC.

Inconsistent findings from some of the previous studies could be partially attributed to the dichotomization of patient population with a single cutoff at the age of 50 years. The assumption of homogeneity of patients older than 50 was not valid as an abundance of literature in geriatric oncology have reported unique treatment response and outcome in very old patients [45, 46]. For example, in the study of Lieu and colleagues, when age was treated as a continuous variable, older age was also associated with worse survival [29], presumably due to reduced life expectancy, increased comorbidities, or inability to tolerate aggressive chemotherapy regimens in older (age>65) patients with mCRC [47]. Therefore, in our study, we divided patients into three age groups (<50, 50-65, and >65 years) for analysis. As expected, we found that patients in both age<50 and age>65 groups had worse OS compared to age 50-65 group. Another distinct feature of our study is that patients in our test cohort for initial analysis received the same treatment, first line FOLFOX for mCRC to minimize possible risk modification from different treatment regimen on survival. Gratifyingly, similar observation has also been found in our real-world patient cohort who received different treatment regimen including singlet, doublet, and triplet chemotherapy with biologics.

Differences in baseline characteristics of patient populations could also prevent us from observing the age-related disparity of survival outcomes in previous studies. In our study, we found a higher proportion of females in patients with EO-mCRC in line with previous studies [24, 27, 28, 42, 48]. The underlying reasons could be due to either a potential selection bias or a differential genetic susceptibility, which warranted further studies. Meanwhile, consistent with previous studies [26, 48], a lower proportion of white patients were observed in EO-m CRC group. Given prior studies that suggested gender and race disparity on survival [49], we performed multivariable analysis to adjust for gender and race in addition to ECOG performance status which was believed to be better at the baseline in younger patients [26].

To utilize excellent recording of treatment-related adverse events in clinical trial data, we also compared them among the three age groups and observed age-related disparity in patients with mCRC. Consistent with previous findings in the adjuvant setting [24-28], our data showed an increased incidence of nausea/vomiting but decreased incidence of diarrhea, fatigue and neutropenia in younger patients. Previously reported potential decrease of toxicity profile in younger patients [27, 50], was not entirely supported by our study since we also observed higher incidence of severe abdominal pain, severe anemia and severe rash in patients with EO-mCRC. In addition, they had earlier onset of nausea/vomiting, mucositis and neutropenia. Hence, it is more appropriate to say that patients with EO-mCRC exhibited unique pattern of adverse events after receiving first-line FOLFOX. Moreover, the association between severe abdominal pain and severe liver toxicity with worse survival in younger patients suggested an individualized approach to the monitoring and management of these unique treatment-related adverse events.

The age-related disparity of survival outcomes and adverse events may suggest a unique underlying disease biology in different age groups. Comprehensive genomic profiling in some studies demonstrated higher prevalence of *TP53* and *CTNNB1* mutations, but lower prevalence of *BRAF* and *APC* mutations in EO-mCRC [11, 28, 43, 51, 52], in line with some of the findings in our study. There was also higher prevalence of *ERBB2* amplification and *CREBBP* mutation in EO-mCRC of our patient cohort. It is still unclear if these differences of genetic alterations might partially explain the observed disparity in survival outcome given again the inconsistent findings across studies and the numerically small differences of a few out of many genes. Thus, further efforts should likely focus on multi-omics studies such as proteogenomic analysis of EO-mCRC to inform disease biology and therapy.

Our study has several strengths over previous studies. First and foremost, we included two additional cohorts, one from another clinical trial, the other one from a real-world patient population, to validate our findings of age-related disparity of survival outcome. Efforts to include external validation cohorts that came from a different time period and have distinct baseline patient characteristics ensured the robustness of our findings. We utilized the excellent quality of prospectively collected clinical trial data on survival endpoints and treatment-related adverse events to study age-related disparity in mCRC. However, these clinical trials were conducted in an era where biologic agents had not become standard first-line therapy for patients with mCRC and clinical NGS was not used to guide treatment. To compensate and address these limitations intrinsic to these data used in our study for analyses, we incorporated our institutional patient and clinical NGS data that were from modern era and more heterogeneous on patients and their treatment as an external validation cohort. In fact, distributions of some important baseline characteristics such as race, tumor sidedness, and first-line treatments in this MCC patient cohort were consistent with those previously reported [26, 28, 31, 48]. However, as a limitation, we did not evaluate adverse events, PFS, and perform multivariable survival analysis in this cohort because retrospective data collection even if some of the data were from prospectively maintained database were very prone to selection bias and inaccuracy.

In conclusion, we observed age-related disparity of survival outcome and treatment-related adverse events in patients with mCRC. Patients with EO-mCRC who received first-line treatment had worse survival compared to their older counterparts and experienced unique treatment-related adverse events. These findings may inform individualized management approaches in patients with early-onset metastatic CRC.

## Data Availability

All data produced in the present study are available upon reasonable request to the authors.

## Notes

### Author contributions

Conceptualization: LM, HX; Methodology: LM, RT, RJ, XW, HX; Data collection: LM, MGD, MFG, TCK, HX; Data analysis and interpretation: LM, RT, RJ, JMH, RDK, HX; Original draft preparation: LM, HX; Manuscript review and editing: All authors.

## Data Availability

The data underlying this article are available in the article and in its online Supplementary Material. The data sharing of individual patient data from each participating trial will be subject to the policy and procedures of the institutions and groups who conducted the original study.

