## Supplemental Tables for "Age-related disparity of survival outcomes and treatment-related adverse events in patients with metastatic colorectal cancer"

**Supplemental table 1.** Demographic and clinical characteristics of patients with metastatic colorectal cancer from all three cohorts.

| | **Study 1** (n=756) | **Study 2** (n=467) | **Moffitt** (n=736) | **P value** | **Overall** (n=1959) |
|---|---|---|---|---|---|
| **Age**, n (%) | | | | <0.001 | |
| < 50 | 115 (15.2) | 64 (13.7) | 196 (26.6) | | 375 (19.1) |
| 50 - 65 | 358 (47.4) | 224 (48.0) | 319 (43.3) | | 901 (46.0) |
| > 65 | 283 (37.4) | 179 (38.3) | 221 (30.0) | | 683 (34.9) |
| **Gender**, n (%) | | | | 0.10 | |
| Female | 304 (40.2) | 182 (39.0) | 328 (44.6) | | 814 (41.6) |
| Male | 452 (59.8) | 285 (61.0) | 408 (55.4) | | 1145 (58.4) |
| **Race**, n (%) | | | | <0.001 | |
| White | 680 (89.9) | 460 (98.5) | 602 (83.3) | | 1742 (89.5) |
| Other | 76 (10.1) | 7 (1.5) | 121 (16.7) | | 204 (10.5) |
| **ECOG**, n (%) | | | | 0.007 | |
| 0/1 | 739 (98.0) | 443 (95.1) | NA | | 1182 (96.9) |
| 2 | 15 (2.0) | 23 (4.9) | NA | | 38 (3.1) |

NA: not available

**Supplemental table 2.** Incidence (%) of adverse events for each age group from Study 1 and Study 2.

| | All grade | | | | Grade 3, 4 & 5 | | | |
|---|---|---|---|---|---|---|---|---|
| AE | Age <50 | Age 50-65 | Age >65 | P | Age <50 | Age 50-65 | Age >65 | P |
| Abdominal pain | 71 (39.7) | 180 (30.9) | 145 (31.4) | 0.082 | 15 (8.4) | 20 (3.4) | 16 (3.5) | 0.018 |
| Anaemia | 23 (12.8) | 53 (9.1) | 45 (9.7) | 0.334 | 11 (6.1) | 6 (1.0) | 7 (1.5) | <0.001 |
| Chest pain | 5 (2.8) | 21 (3.6) | 12 (2.6) | 0.656 | 0 (0.0) | 1 (0.2) | 1 (0.2) | 0.990 |
| Diarrhoea | 92 (51.4) | 281 (48.3) | 264 (57.1) | 0.017 | 11 (6.1) | 53 (9.1) | 60 (13.0) | 0.020 |
| DVT/PE | 8 (4.5) | 33 (5.7) | 35 (7.6) | 0.286 | 4 (2.2) | 23 (4.0) | 26 (5.6) | 0.152 |
| Fatigue | 79 (44.1) | 273 (46.9) | 257 (55.6) | 0.0052 | 8 (4.5) | 32 (5.5) | 44 (9.5) | 0.019 |
| Liver toxicity | 9 (5.0) | 31 (5.3) | 14 (3.0) | 0.179 | 3 (1.7) | 8 (1.4) | 8 (1.7) | 0.858 |
| Mucositis | 66 (36.9) | 183 (31.4) | 164 (35.5) | 0.242 | 4 (2.2) | 6 (1.0) | 15 (3.2) | 0.034 |
| Nausea/vomiting | 124 (69.3) | 335 (57.6) | 279 (60.4) | 0.019 | 5 (2.8) | 20 (3.4) | 23 (5.0) | 0.346 |
| Neuropathy | 153 (85.5) | 494 (84.9) | 396 (85.7) | 0.932 | 32 (17.9) | 99 (17.0) | 84 (18.2) | 0.881 |
| Neutropenia | 69 (38.5) | 231 (39.7) | 230 (49.8) | 0.0018 | 46 (25.7) | 154 (26.5) | 176 (38.1) | <0.001 |
| Rash | 52 (29.1) | 143 (24.6) | 104 (22.5) | 0.224 | 5 (2.8) | 7 (1.2) | 2 (0.4) | 0.047 |
| Thrombocytopenia | 31 (17.3) | 123 (21.1) | 117 (25.3) | 0.068 | 4 (2.2) | 13 (2.2) | 19 (4.1) | 0.195 |

**Supplemental table 3.** Median (IQR) time to onset (weeks) of adverse events for each age group from Study 1 and Study 2. IQR, interquartile range.

| | < 50<br>N=141 | 50-65<br>N=446 | > 65<br>N=362 | P |
|---|---|---|---|---|
| Fatigue | 4.1 [1.3;11.6] | 5.4 [1.8;14.1] | 4.6 [1.0;14.4] | 0.356 |
| Nausea/vomiting | 1.0 [0.4;5.0] | 2.1 [0.4;6.4] | 2.6 [0.6;9.7] | 0.012 |
| Diarrhoea | 4.3 [1.6;11.6] | 4.6 [1.7;11.9] | 3.4 [0.9;10.0] | 0.044 |
| Mucositis | 3.6 [0.9;8.8] | 5.1 [2.0;12.1] | 5.7 [2.1;13.9] | 0.051 |
| Neuropathy | 4.3 [0.9;12.3] | 4.8 [2.1;14.4] | 4.6 [2.0;14.4] | 0.697 |
| Fatigue | 4.1 [1.3;11.6] | 5.4 [1.8;14.1] | 4.6 [1.0;14.4] | 0.356 |
| Neutropenia | 8.0 [4.0;14.4] | 9.4 [6.1;15.2] | 8.4 [4.1;14.1] | 0.043 |
| Thrombocytopenia | 12.6 [7.1;17.9] | 12.4 [7.9;18.5] | 12.0 [6.3;21.1] | 0.993 |
| Chest pain | 8.1 [2.3;13.6] | 18.0 [7.1;30.4] | 12.0 [4.5;16.5] | 0.195 |
| DVT/PE | 14.1 [7.3;23.2] | 8.1 [4.3;15.3] | 8.1 [5.4;14.6] | 0.366 |
| Anaemia | 4.7 [2.2;15.3] | 8.1 [4.1;16.6] | 10.1 [2.6;20.1] | 0.535 |
| Liver toxicity | 10.1 [8.3;17.3] | 19.5 [6.1;25.8] | 8.5 [7.0;16.5] | 0.521 |

**Supplemental table 4.** Median (IQR) time to resolution (weeks) of adverse events for each age group from Study 1 and Study 2. IQR, interquartile range.

| | < 50<br>N=112 | 50-65<br>N=305 | > 65<br>N=246 | P |
|---|---|---|---|---|
| Fatigue | 1.0 [0.4;2.0] | 0.9 [0.3;2.7] | 1.0 [0.3;2.4] | 0.931 |
| Nausea/vomiting | 0.3 [0.1;0.6] | 0.3 [0.0;0.6] | 0.3 [0.1;0.7] | 0.240 |
| Diarrhoea | 0.3 [0.1;0.6] | 0.3 [0.1;0.7] | 0.3 [0.1;0.7] | 0.642 |
| Mucositis | 0.6 [0.3;1.0] | 0.9 [0.3;1.9] | 1.0 [0.6;1.6] | 0.006 |
| Neuropathy | 0.6 [0.3;2.0] | 0.7 [0.3;1.9] | 0.7 [0.4;1.7] | 0.937 |
| Fatigue | 1.0 [0.4;2.0] | 0.9 [0.3;2.7] | 1.0 [0.3;2.4] | 0.931 |
| Neutropenia | 1.0 [0.9;1.9] | 1.0 [0.9;1.4] | 1.0 [1.0;1.9] | 0.060 |
| Thrombocytopenia | 1.1 [0.9;2.1] | 1.0 [1.0;2.2] | 1.0 [1.0;2.9] | 0.850 |
| Chest pain | 0.6 [0.0;6.6] | 0.2 [0.0;0.4] | 1.9 [0.0;3.7] | 0.401 |
| DVT/PE | 1.3 [0.4;4.7] | 6.6 [2.0;14.2] | 5.7 [0.9;11.1] | 0.379 |
| Anaemia | 3.3 [0.6;5.2] | 2.0 [0.9;5.0] | 1.3 [0.2;3.8] | 0.471 |
| Liver toxicity | 1.6 [1.0;2.2] | 2.1 [1.2;4.3] | 2.0 [1.6;2.7] | 0.483 |

**Supplemental table 5.** The prognostic value of adverse events (none vs grade 1-2 vs grade 3-5) in patients with early-onset colorectal cancer from Study 1 and Study 2.

| | | Overall survival | | | Progression free survival | | |
|---|---|---|---|---|---|---|---|
| AE | Grade | HR (95% CI) | P Value | Overall P Value | HR (95% CI) | P Value | Overall P Value |
| Abdominal pain | None | 1.0 (Reference) | - | 0.015 | 1.0 (Reference) | - | 0.006 |
| | 1-2 | 1.46 (0.98, 2.18) | 0.066 | | 1.19 (0.84, 1.68) | 0.323 | |
| | 3-5 | 2.24 (1.23, 4.09) | 0.008 | | 2.51 (1.43, 4.41) | 0.001 | |
| Anaemia | None | 1.0 (Reference) | - | 0.664 | 1.0 (Reference) | - | 0.682 |
| | 1-2 | 1.34 (0.68, 2.65) | 0.401 | | 1.30 (0.72, 2.36) | 0.381 | |
| | 3-5 | 1.17 (0.54, 2.52) | 0.692 | | 1.03 (0.52, 2.04) | 0.924 | |
| Chest pain | None | 1.0 (Reference) | - | | 1.0 (Reference) | - | |
| | 1-2 | 2.33 (0.85, 6.34) | 0.099 | | 1.50 (0.61, 3.67) | 0.375 | |
| Diarrhoea | None | 1.0 (Reference) | - | 0.528 | 1.0 (Reference) | - | 0.528 |
| | 1-2 | 0.83 (0.57, 1.21) | 0.342 | | 0.83 (0.60, 1.15) | 0.259 | |
| | 3-5 | 1.19 (0.51, 2.77) | 0.690 | | 0.91 (0.47, 1.76) | 0.773 | |
| DVT/PE | None | 1.0 (Reference) | - | 0.986 | 1.0 (Reference) | - | 0.419 |
| | 1-2 | 1.06 (0.33, 3.34) | 0.927 | | 1.58 (0.58, 4.29) | 0.370 | |
| | 3-5 | 1.09 (0.34, 3.43) | 0.886 | | 0.62 (0.23, 1.67) | 0.343 | |
| Fatigue | None | 1.0 (Reference) | - | 0.099 | 1.0 (Reference) | - | 0.969 |
| | 1-2 | 0.66 (0.44, 0.97) | 0.035 | | 1.01 (0.72, 1.39) | 0.976 | |
| | 3-5 | 0.71 (0.31, 1.64) | 0.421 | | 1.10 (0.53, 2.27) | 0.801 | |
| Liver toxicity | None | 1.0 (Reference) | - | 0.104 | 1.0 (Reference) | - | 0.104 |
| | 1-2 | 0.62 (0.23, 1.68) | 0.344 | | 0.59 (0.24, 1.44) | 0.247 | |
| | 3-5 | 3.99 (0.95, 16.76) | 0.059 | | 2.82 (0.89, 8.97) | 0.079 | |
| Mucositis | None | 1.0 (Reference) | - | 0.098 | 1.0 (Reference) | - | 0.588 |
| | 1-2 | 0.64 (0.43, 0.97) | 0.035 | | 0.90 (0.64, 1.25) | 0.523 | |
| | 3-5 | 1.13 (0.35, 3.57) | 0.841 | | 1.45 (0.53, 3.96) | 0.466 | |
| Nausea/vomiting | None | 1.0 (Reference) | - | 0.597 | 1.0 (Reference) | - | 0.262 |
| | 1-2 | 0.98 (0.66, 1.45) | 0.902 | | 0.91 (0.64, 1.28) | 0.587 | |
| | 3-5 | 1.78 (0.54, 5.82) | 0.340 | | 1.91 (0.75, 4.86) | 0.172 | |
| Neuropathy | None | 1.0 (Reference) | - | 0.251 | 1.0 (Reference) | - | 0.005 |
| | 1-2 | 0.85 (0.54, 1.34) | 0.484 | | 0.68 (0.46, 1.01) | 0.055 | |
| | 3-5 | 0.60 (0.32, 1.10) | 0.100 | | 0.43 (0.26, 0.71) | 0.001 | |
| Neutropenia | None | 1.0 (Reference) | - | 0.884 | 1.0 (Reference) | - | 0.743 |
| | 1-2 | 0.93 (0.53, 1.62) | 0.792 | | 0.97 (0.60, 1.57) | 0.902 | |
| | 3-5 | 0.90 (0.59, 1.38) | 0.638 | | 0.87 (0.60, 1.25) | 0.442 | |
| Rash | None | 1.0 (Reference) | - | 0.141 | 1.0 (Reference) | - | 0.590 |
| | 1-2 | 0.63 (0.40, 1.00) | 0.049 | | 0.83 (0.57, 1.19) | 0.304 | |
| | 3-5 | 0.81 (0.26, 2.57) | 0.723 | | 0.94 (0.38, 2.31) | 0.891 | |
| Thrombocytopenia | None | 1.0 (Reference) | - | 0.671 | 1.0 (Reference) | - | 0.572 |
| | 1-2 | 0.83 (0.49, 1.41) | 0.488 | | 1.15 (0.74, 1.79) | 0.522 | |
| | 3-5 | 1.30 (0.48, 3.55) | 0.610 | | 1.57 (0.58, 4.26) | 0.377 | |

**Supplemental table 6.** Demographic and clinical characteristics of patients with metastatic colorectal cancer in the Moffitt Cancer Center cohort stratified by age groups.

| | **< 50** (n=196) | **50 - 65** (n=319) | **> 65** (n=221) | **P value** | **Overall** (n=736) |
|---|---|---|---|---|---|
| **Gender**, n (%) | | | | 0.49 | |
| Female | 88 (44.9) | 135 (42.3) | 105 (47.5) | | 328 (44.6) |
| Male | 108 (55.1) | 184 (57.7) | 116 (52.5) | | 408 (55.4) |
| **Race**, n (%) | | | | 0.044 | |
| White | 159 (81.1) | 255 (79.9) | 188 (85.5) | | 602 (81.8) |
| Other | 37 (18.9) | 64 (20.1) | 33 (14.5) | | 134 (18.2) |
| **First-line therapy**, n (%) | | | | <0.001 | |
| Single | 12 (6.1) | 17 (5.3) | 32 (14.5) | | 61 (8.3) |
| Doublet | 157 (80.1) | 283 (88.7) | 184 (83.3) | | 624 (84.8) |
| Triplet | 27 (13.8) | 19 (6.0) | 5 (2.3) | | 51 (6.9) |
| **Sidedness**, n (%) | | | | | |
| Left | 142 (76.3) | 221 (71.1) | 118 (57.6) | <0.001 | 481 (68.5) |
| Right | 44 (23.7) | 90 (28.9) | 87 (42.4) | | 221 (31.5) |
| **MSI status**, n (%) | | | | 0.29 | |
| MSI | 8 (4.4) | 16 (5.7) | 16 (8.2) | | 40 (6.1) |
| MSS | 172 (95.6) | 265 (94.3) | 178 (91.8) | | 615 (93.9) |

**Supplemental table 7.** Prevalence of common gene mutations in patients with metastatic colorectal cancer from the Moffitt Cancer Center cohort stratified by age groups.

| **Gene** | **<50** (n=196) n (%) | **50-65** (n=319) n (%) | **>65** (n=221) n (%) | **P Value** |
|---|---|---|---|---|
| *TP53* | 153 (78.1) | 226 (70.8) | 155 (70.1) | 0.13 |
| *APC* | 154 (78.6) | 243 (76.2) | 158 (71.5) | 0.22 |
| *KRAS* | 92 (46.9) | 151 (47.3) | 105 (47.5) | 0.99 |
| *PIK3CA* | 30 (15.3) | 63 (19.7) | 40 (18.1) | 0.44 |
| *SMAD4* | 30 (15.3) | 41 (12.9) | 35 (15.8) | 0.57 |
| *BRAF* | 15 (7.7) | 27 (8.5) | 37 (16.7) | 0.002 |
| *FBXW7* | 15 (7.7) | 32 (10.0) | 19 (8.6) | 0.64 |
| *ARID1A* | 15 (7.7) | 21 (6.6) | 19 (8.6) | 0.68 |
| *CTNNB1* | 13 (6.6) | 10 (3.1) | 5 (2.3) | 0.047 |
| *ATM* | 12 (6.1) | 18 (5.6) | 15 (6.8) | 0.86 |
| *EGFR* | 11 (5.6) | 14 (4.4) | 9 (4.1) | 0.73 |
| *ERBB2*.amp | 10 (5.1) | 2 (0.6) | 5 (2.3) | 0.005 |
| *NRAS* | 8 (4.1) | 15 (4.7) | 5 (2.3) | 0.34 |
| *PTEN* | 7 (3.6) | 23 (7.2) | 11 (5.0) | 0.20 |
| *MYC*.amp | 7 (3.6) | 13 (4.1) | 10 (4.5) | 0.89 |
| *NF1* | 6 (3.1) | 12 (3.8) | 5 (2.3) | 0.62 |
| *CREBBP* | 6 (3.1) | 3 (0.9) | 1 (0.5) | 0.050 |
